# Massively parallel functional profiling identifies *CCDC88C* as a risk gene for ER-positive breast cancer

**DOI:** 10.64898/2026.03.02.26347419

**Authors:** Kate Mackie, Harriet Kemp, Andrea Gunnell, James B. Studd, Molly Went, Philip Law, Katarzyna Tomczyk, Selin Sevgi, Yunjiao Lu, Nick Orr, Richard S. Houlston, Nichola Johnson, Olivia Fletcher, Syed Haider

## Abstract

Genome wide association studies (GWAS), combined with fine-mapping have identified 196 independent signals associated with breast cancer risk. Deciphering the functional basis of these associations can inform our understanding of the biology and aetiology of breast cancer. Decoding GWAS risk associations is challenging due to linkage disequilibrium between variants and because most variants map to non-coding regions, influencing breast cancer risk via cis-regulatory mechanisms that modulate the expression of target genes. To identify the functional variants driving breast cancer risk associations, we carried out a lentivirus-based massively parallel reporter assay (lentiMPRA) to screen 5,116 credible causal variants across these signals. We identified 709 variants mapping to 140 risk regions, that are associated with significant variation between REF and ALT alleles. A follow-up investigation at 14q32.11 revealed rs7153397 may impact expression of *CCDC88C* to influence both breast cancer risk and prognosis. These findings provide a prioritised set of functional variants for downstream analyses, advancing our understanding of breast cancer risk mechanisms.

## MAIN

Breast cancer, which affects over 2 million women worldwide annually, has a strong heritable basis. Genome-wide association studies (GWAS) of breast cancer, combined with fine-mapping, have identified 196 independent signals associated with breast cancer risk^1^. Deciphering the functional basis of these risk associations has the potential to inform on the biology and aetiology of breast cancer. However, decoding GWAS risk associations is challenging due to linkage disequilibrium (LD) between variants and because most variants localise to non-coding regions.

Fine-mapping analysis, by the Breast Cancer Association Consortium (BCAC) and the Consortium of Investigators of Modifiers of BRCA1/2 (CIMBA), reported 7,394 credible causal variants (CCVs) as the functional basis of these 196 associations^1^. CCVs are defined as variants with an association *P*-value within two orders of magnitude of an index variant^2^. While such computational approaches can prioritize variants and predict candidate target genes using *in silico* data, experimental validation is required to confirm regulatory activity and target genes. Most non-coding GWAS risk variants are thought to act via cis-regulatory mechanisms that modulate the expression of their target genes. By examining transcriptional changes linked to genetic variants, specific alleles can be associated with altered gene expression.

While classical reporter assays are limited to assessing the allelic activity of individual variants, massively parallel reporter assays (MPRAs) offer a scalable approach to evaluate the regulatory effects of thousands of variants simultaneously. To systematically assay the allele-specific regulatory activity associated with variants at breast cancer risk loci we carried out a lentivirus-based massively parallel reporter assay (lentiMPRA)^3^ to screen 5,116 of the BCAC reported CCVs across 195 of the independent risk signals (1 to 351 CCVs per signal)^1^.

Here we report the results of this lentiMPRA analysis in T-47D breast cancer cells. We identify a total of 709 credible causal variants (CCVs) across 140 risk regions that exhibit significant allelic differences in regulatory activity, supporting their role as functional drivers of genetic association. Follow-up analysis of rs7153397 at 14q32.11 identifies *CCDC88C* as the target gene at this locus, with its expression associated with breast cancer subtype and survival in estrogen receptor positive (ER+) cases.

To determine which CCVs reported by BCAC^1^ are functional, we adapted a lentiMPRA to screen 5,116 CCVs across 195 independent risk signals in the breast cancer cell line T-47D. We generated a plasmid library of 20,878 oligonucleotides, each 270 bp centred on the reference (REF) or alternative (ALT) alleles in forward and reverse orientations, linked to unique barcodes (**Figure 1A**). The library was packaged into lentivirus and used to infect T-47D breast cancer cells in triplicate, and sequenced to assess regulatory activity via barcode transcription^3^ (**Figure 1B**). T-47D cells were chosen for their estrogen receptor-positive (ER+) profile and availability of epigenetic (DNase I hypersensitive sites (DHS), H3K27ac and transcription factor (TF) ChIP-seq) data.

**Figure 1.**
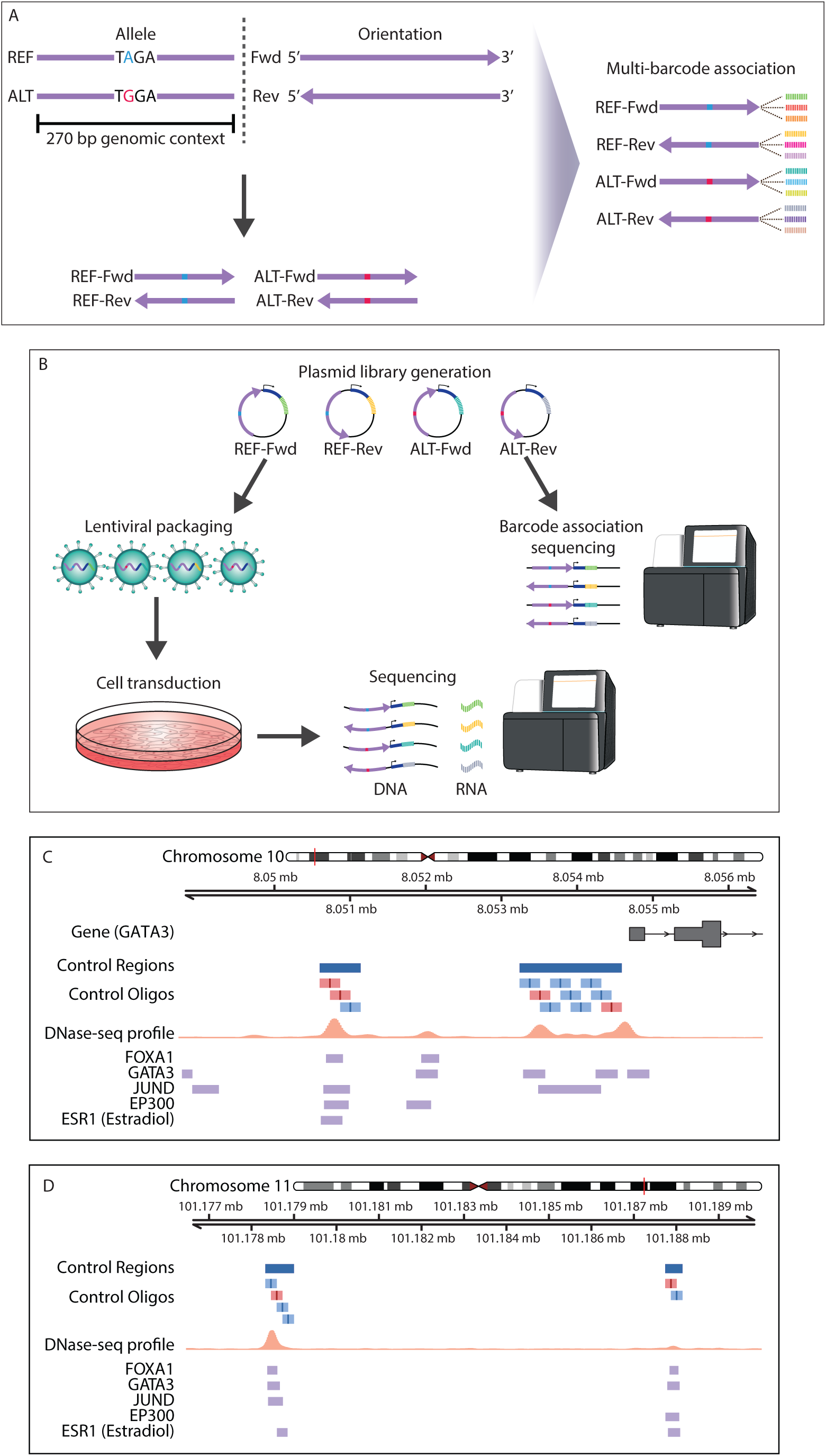
LentiMPRA study design. **A,** Oligonucleotides containing reference (REF) or alternative (ALT) allele and in forward (Fwd) or reverse (Rev) orientation were linked to unique barcodes to generate an oligonucleotide library. **B.** Barcoded oligonucleotides were inserted into plasmids, and packaged into lentivirus. T-47D cells were infected, and DNA / RNA sequencing used to assess regulatory activity. **C and D**, Genomic regions near *GATA3* (**C**) and *PGR* (**D**) with oligonucleotides coloured according to enhancer activity (red: significant, blue: non-significant) and T-47D annotations. DNase sequencing profile, orange; TF ChIP-seq, purple.

As positive controls, we designed oligonucleotides targeting regions near *GATA3* and *PGR* (**Figure 1C, D**), known for their regulatory roles in ER+ breast cancer^4,5^. These comprised 36 oligonucleotides across regions with DHS and TF binding (*FOXA1*, *GATA3*, *JUND*, *EP300*, *ESR1*; **Figure 1C, D**). Negative controls comprised 400 oligonucleotides, from 100 scrambled sequences. The library contained 3.64 million unique barcodes, covering 99.16% of designed oligonucleotides (mean 175 barcodes per oligonucleotide). Three replicates showed strong correlation (r > 0.97; **Supplementary Figure 1**).

We used MPRAnalyze^6^ to quantify enhancer activity (alpha) and differential activity between alleles. After filtering oligonucleotides with < 20 or > 200 barcodes, we analysed 4,732 (92.49%) test CCVs, 17 (94.44%) positive controls and 99 (99%) negative controls. Among controls, 35.29% of positive controls and 15.5% of negative controls showed significant enhancer activity (alpha, FDR-adjusted *P* < 0.1). For test CCVs, 6.47% (306/4732) showed significant enhancer activity, implying these CCVs were mapping to regulatory sequences (**Figure 2A**).

**Figure 2.**
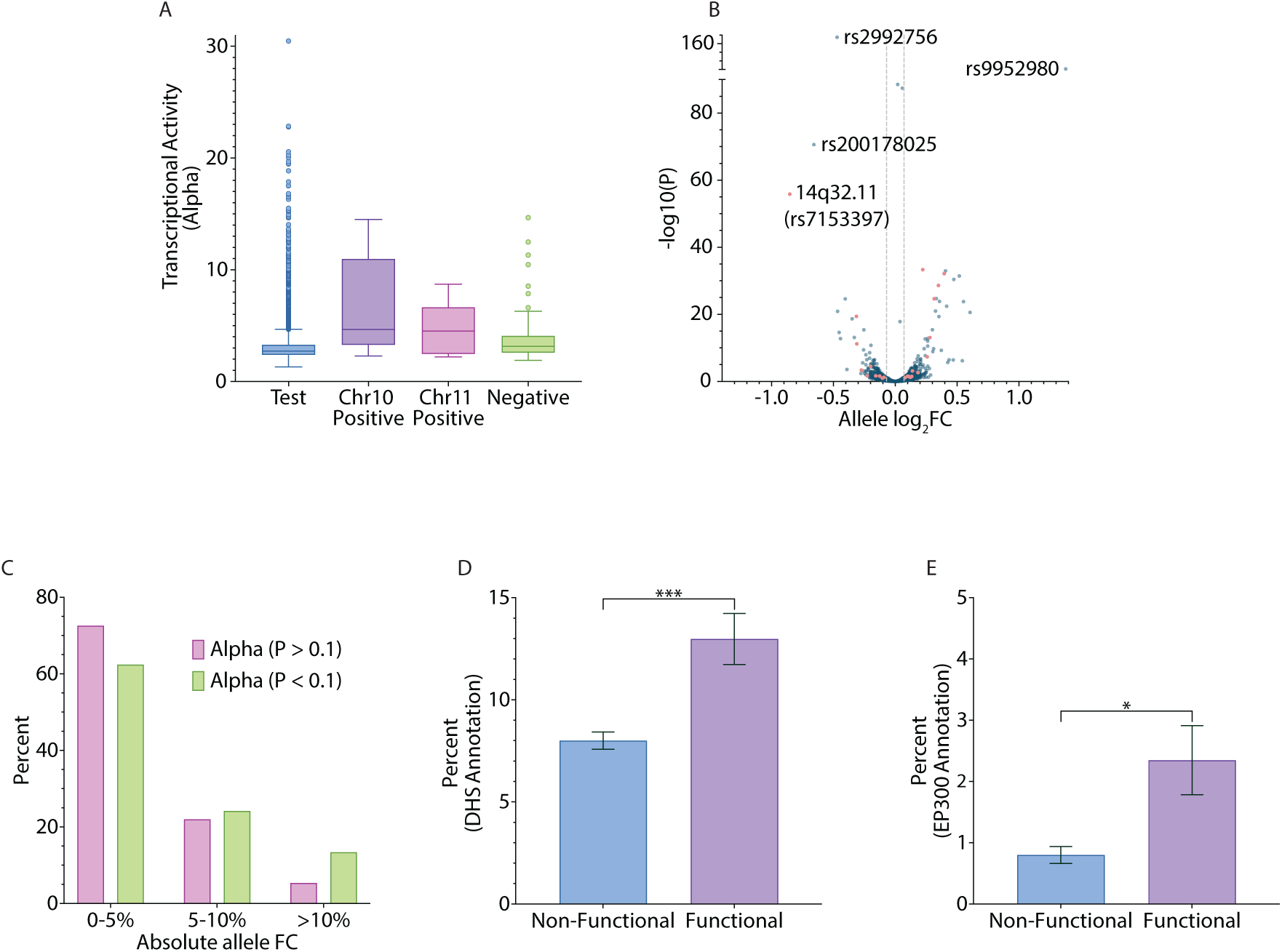
LentiMPRA identifies functional breast cancer associated risk variants. **A,** Boxplot of enhancer activity for test, positive and negative control sequences. The box represents the interquartile range (IQR), with whiskers 1.5 x IQR, the centre line represents the median. **B,** Volcano plot of allele log_2_FC vs -log10 of FDR-corrected *P* for test sequences, with vertical lines at +/-0.07 log_2_FC (equivalent to +/-5% difference in activity between alleles). Red dots indicate 23 variants prioritised for follow-up. **C,** Proportion of variants with increasing absolute allele FC, stratified by enhancer activity *P* = 1.51 ×10^-8^. **D, E,** Percentage of sequences with DHS (D) or *EP300* (E) annotations comparing non-functional (allele log_2_FC *P* > 0.1) and functional variants (allele log_2_FC *P* < 0.1). * *P* < 0.006, *** *P* < 0.00006.

We identified 709 CCVs (14.98%) at 140 risk signals which showed significant differential activity between alleles (FDR-adjusted *P* < 0.1), ranging from -0.85 to 1.38 log_2_ fold change (FC) (**Figure 2B, Supplementary Data**). Most of these (93.65%), showed modest effects (< 20% differential activity) consistent with common variants individually making a small contribution to overall risk. On average there were five CCVs showing differential allelic activity per risk signal (range 1-42). Stratified analysis confirmed that allele log_2_FC was consistent across forward and reverse orientation (**Supplementary Figure 2; Supplementary Table 1**).

CCVs showing significant enhancer activity were enriched for larger allelic differences (|allele FC| > 10%, *P* = 1.51×10^-8^; **Figure 2C**). These variants also showed significant enrichment for DHS (*P* = 3.19×10^-5^) and EP300 binding (*P* = 8.79×10^-4^) in T-47D cells (**Figure 2D, E**) supporting their regulatory impact (**Supplementary Figure 3**).

Comparing the 709 differentially expressed variants to BCAC data, 5.4% were index variants (1.5-fold enrichment, *P* = 0.03) and 1.0% were single CCVs (2-fold enrichment, *P* = 0.11). We also validated six variants which are the subject of single-locus studies (**Supplementary Table 2**) and noted four additional variants that displayed strong enhancer activity but no allelic differences, making it possible that some previous studies prioritised enhancer presence over functional impact *per se*.

Variant prioritisation was performed by filtering for those with |log₂FC| > 0.07, excluding promoter regions, and selecting variants with significant enhancer activity and relevant epigenetic marks (**Figure 3A, Supplementary Table 3**; full dataset **Supplementary Data**). Amongst the 23 prioritised candidates, rs7153397 at 14q32.11 exhibited a log₂FC of 0.85 (80% increase in activity; *P* = 1.5×10⁻⁵⁶) and was selected for further analysis. This variant is in perfect LD with the regional index variant rs11341843 (D′ = 1.0, r² = 1.0; odds ratio for ER-positive breast cancer = 1.06, *P* = 1.4×10⁻¹¹)^1^, which did not have a significant allelic difference (*P* = 0.53). rs7153397 also maps to a region marked by H3K27ac and binding of FOXA1, ESR1, and EP300, consistent with a putative regulatory mechanism underlying increased breast cancer risk (**Figure 3B, C**).

**Figure 3.**
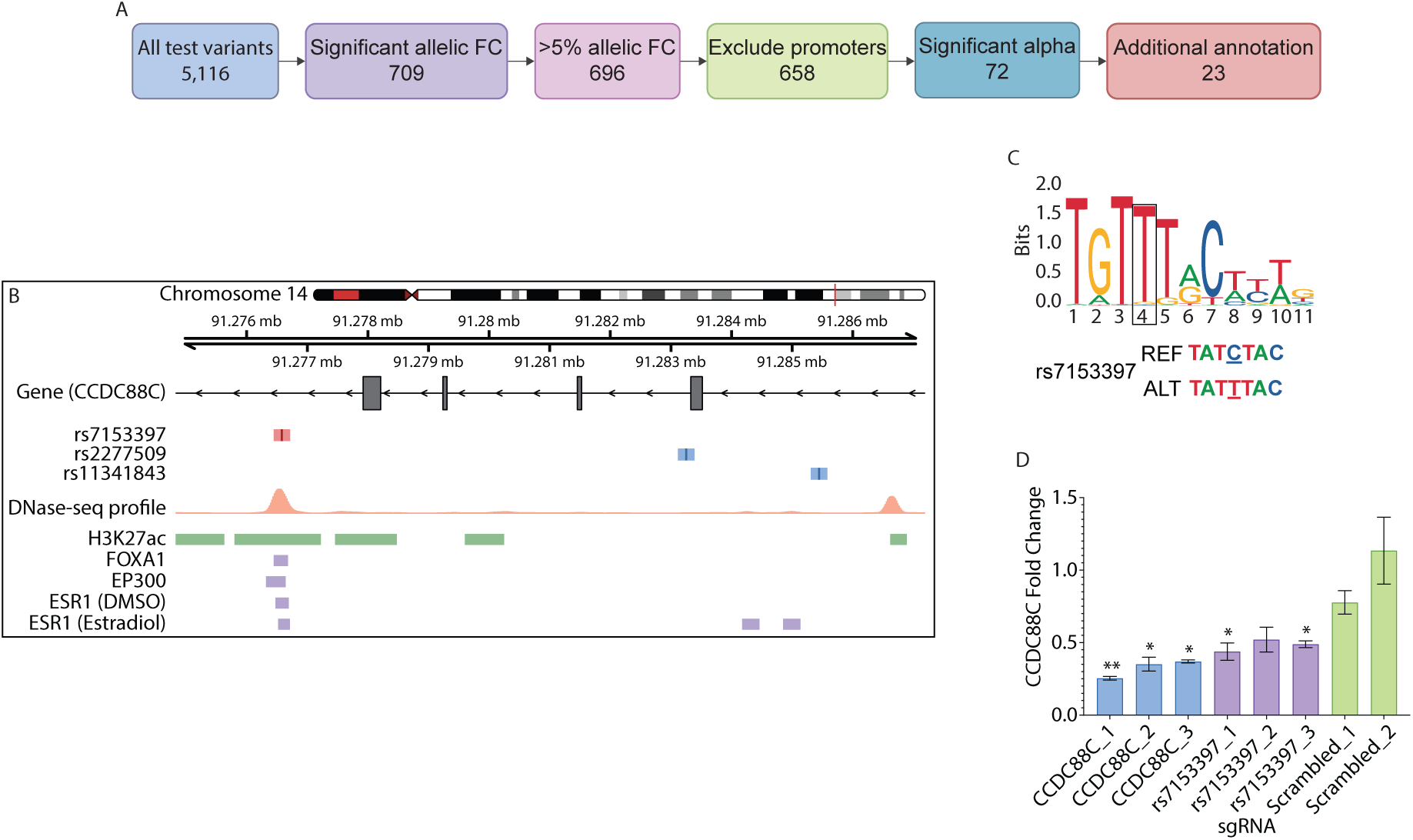
*CCDC88C* as a target gene for rs7153397 at 14q32.11. **A,** Pipeline to selecting rs7153397 for functional follow up. **B,** Genomic region at 14q32.11 with test sequences (red: rs7153397, blue: other variants) and T-47D annotations. DNase sequencing profile, orange; H3K27ac, green; TF ChIP-seq, purple. **C,** FOXA1 binding matrix with rs7153397 reference (REF) and alternative (ALT) sequences of rs7153397 shown below; the base corresponding to position 4 is underlined. **D**, *CCDC88C* expression levels in T-47D cells following CRISPRi with sgRNAs targeting dCas9-KRAB to the *CCDC88C* promoter (CCDC88C_1, CCDC88C_2, CCDC88C_3), rs7153397 variant (rs7153397_1, rs7153397_2, rs7153397_3) or non-targeting negative controls (Scrambled_1, Scrambled_2). Fold change was corrected for *GAPDH* and relative expression (compared to empty vector alone) was calculated for each sgRNA using the ΔΔCT method. Error bars correspond to standard error of the mean; *P* values from *t*-tests. A Bonferroni *P* value of 0.006 was considered significant. * *P* < 0.006, ** *P* < 0.0006.

To functionally validate the regulatory role of rs7153397 and its putative target gene, *CCDC88C*, CRISPR interference (CRISPRi) was performed in T-47D cells using dCas9-KRAB and sgRNAs targeting the variant. Two rs7153397-specific sgRNAs significantly reduced *CCDC88C* expression (*P* = 0.002 and 0.004), with no effect on neighbouring genes (*C14orf1159* or *PPP4R3A*; **Supplementary Figure 4**), confirming *CCDC88C* as the likely target gene underlying the rs7153397–14q32.11 association (**Figure 3D**). In the Cancer Genome Atlas (TCGA), *CCDC88C* expression was significantly elevated in ER+ and ER-breast tumours compared to normal breast tissue (*P* = 2.0×10⁻²³ and *P* = 0.0002, respectively) and in both TCGA and the Sweden Cancerome Analysis Network-Breast (SCAN-B) cohorts, *CCDC88C* expression was significantly higher in ER+ compared to ER-tumours (*P* = 2.2×10⁻¹⁵ in TCGA; *P* = 1.4×10⁻³⁰ in SCAN-B) (**Figure 4A, B**). Higher *CCDC88C* expression was also associated with improved survival in ER+ disease (hazard ratio (HR) = 0.53, *P* = 0.004 in TCGA; HR = 0.63, *P* = 1.9×10⁻⁵ in SCAN-B; **Figure 4C, D**) but not ER-disease (**Figure 4e, F**).

**Figure 4.**
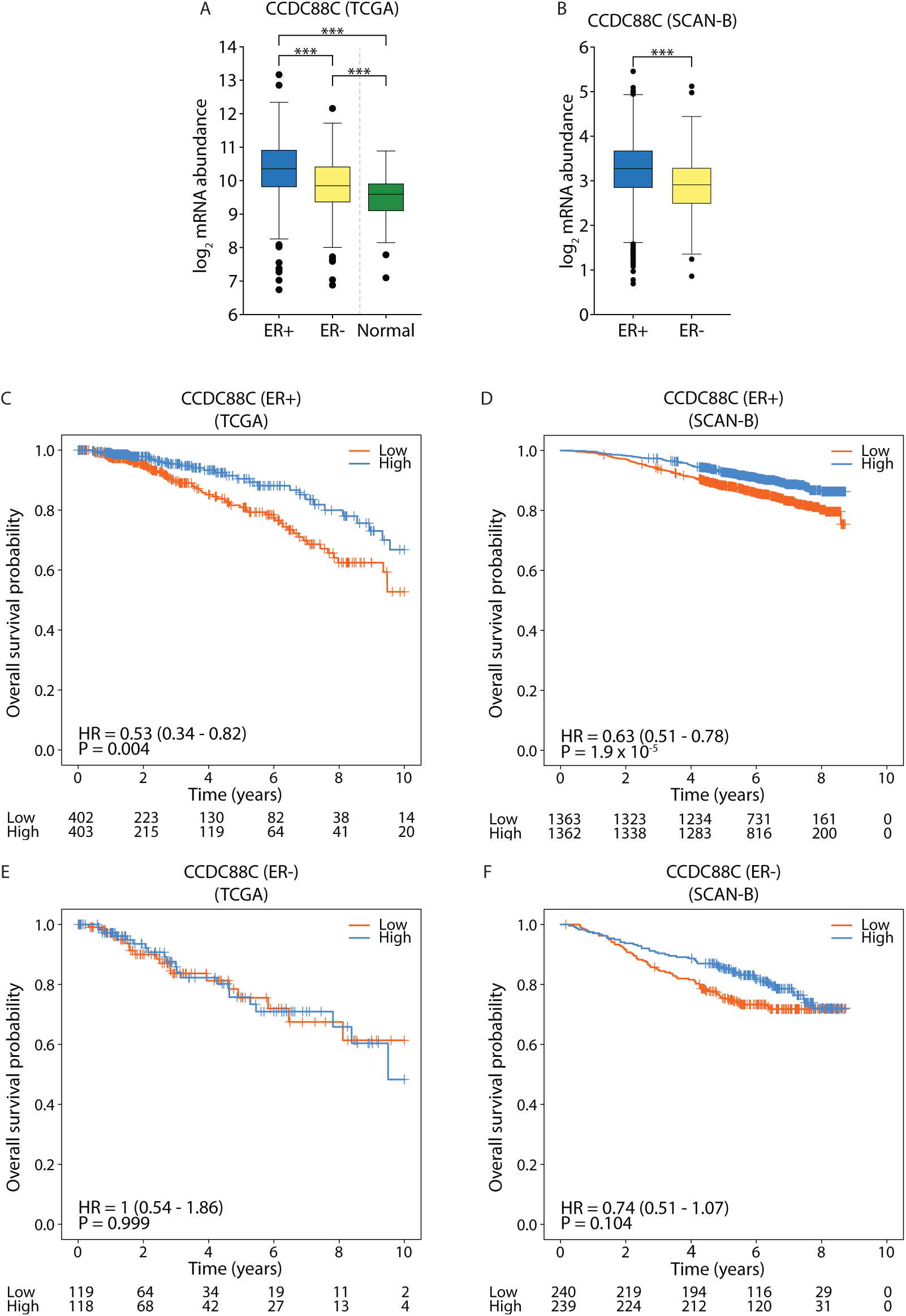
CCDC88C expression is associated with breast cancer subtype and survival. **A, B,** Boxplots of log_2_ abundance of *CCDC88C* mRNA in breast tissue stratified by ER status in data from **(A)** TCGA and **(B)** SCAN-B; * *P* < 0.01, ** *P* < 0.001, *** *P* < 0.0001. The box represents the IQR, with whiskers 1.5 x IQR, the centre line represents the median. **C-F,** Kaplan-Meier plots of association between *CCDC88C* expression (above median expression (high) versus below median expression (low)) in ER+ data from TCGA **(A)** and SCAN-B **(B)**, and ER-data from TCGA **(C)** and SCAN-B **(D)**.

We report the use of a high-throughput barcoded lentiMPRA to assess the functional effect of 5,116 CCVs identified by BCAC fine-mapping analysis^1^. We identified 709 CCVs across 140 risk regions which show statistically significant allelic effects on reporter activity. Among these, we characterised rs7153397 at 14q32.11, identifying *CCDC88C* as its target gene and proposing a mechanism by which this variant might alter expression of *CCDC88C* to impact breast cancer risk.

Our choice of barcoded lentiMPRA to assay multiple variants was driven by its advantages over alternatives, such as STARR-seq. Although STARR-seq offers simpler protocols and supports longer sequences^7^, lentiMPRA’s use of multiple unique barcodes per candidate regulatory sequence (CRS) minimizes biases from RNA stability, thereby enabling more precise quantification of enhancer activity^8^. Furthermore, lentiMPRA’s integration of reporter constructs into the host cell genome better recapitulates the *in vivo* chromatin context when compared to episomal assays, such as STARR-seq^9,10^. These features thereby maximise the reliability of our functional readouts, especially for assessing regulatory activity in a chromatin-relevant setting.

Our study represents the first comprehensive functional validation of BCAC-defined CCVs, which were identified using a combination of stepwise multinomial logistic regression and evaluated using the PAINTOR Bayesian model, which integrates genetic association, LD, and epigenomic data^1^. Overall, we observed only a weak correlation between PAINTOR posterior probabilities (PP) and both allelic activity (log₂FC; Spearman’s *ρ* = 0.12) and enhancer strength (alpha; Spearman’s *ρ* = 0.08). However, among the eight variants predicted as being causal for ER+ breast cancer (PP > 0.8), five showed a significant difference in allelic activity, including rs7153397, with two also exhibiting enhancer activity. While four variants have been the subject of previous locus-specific analysis, the identification of rs7153397 as the functional basis of the 14q23 association represents a novel finding. Our findings therefore serve to highlight the complementary value of fine-mapping, high-throughput screening, and targeted follow-up in prioritizing causal variants.

Despite the enrichment of variants with significant allelic effects in strong enhancers (FDR-adjusted *P* < 0.1), overlap was incomplete. We identified 222 CCVs lacking allelic differences that mapped to strong enhancers, suggesting these variants reside in regulatory regions but do not directly modulate enhancer activity, including four previously validated functional variants mapping to strong enhancers but lacking allelic effects. This discrepancy underscores the limitations of relying solely on epigenomic annotations, which may prioritize variants in active regulatory regions without confirming their functional impact. Additionally, 625 variants with significant allelic effects mapped to sequences without detectable enhancer activity. We acknowledge that this may reflect technical constraints, as the 270 bp oligonucleotides used may not fully capture the regulatory context of enhancers^11^. Hence these variants may reside in weak or context-dependent enhancers or require specific cellular or environmental cues for activity.

Our follow-up analysis of rs7153397 identified *CCDC88C* as the likely target gene at the 14q32.11 locus. *CCDC88C* encodes a regulator of non-canonical Wnt signalling via the PI3K–AKT pathway^12^, which plays a role in mammary gland development^13^ and breast cancer progression^14^, but to date has not been implicated in early tumorigenesis. We demonstrated that rs7153397 resides within a putative FOXA1 binding site, and CRISPRi-mediated targeting of this site reduced *CCDC88C* expression, supporting a regulatory mechanism underlying its association with breast cancer risk. Consistent with the link between rs7153397 and ER+ breast cancer, *CCDC88C* expression was consistently increased in ER+ tumours with higher expression being associated with better prognosis. These data support *CCDC88C* upregulation contributing to ER+ breast cancer and influencing patient prognosis.

In conclusion, our study identifies a functionally annotated subset of breast cancer risk variants that modulate enhancer activity, providing a refined list of high-priority CCVs for mechanistic follow-up. Unlike epigenomic annotations alone, our approach pinpoints variants with demonstrable regulatory effects, offering deeper insight into the functional architecture of breast cancer susceptibility.

## METHODS

### Experimental design

We screened 5,116 breast cancer associated credible causal variants (CCVs), which were reported by the Breast Cancer Association Consortium (BCAC) fine mapping analysis^1^, using a lentivirus-based massive parallel reporter assay (lentiMPRA)^3^ in T-47D cells to identify variants with differential regulatory activity. We designed 270 bp oligonucleotides centred on reference (REF) or alternative (ALT) alleles, packaged them into a lentivirus library and infected T-47D cells. We sequenced DNA and RNA to assess regulatory activity, using available datasets to annotate our library. We followed up on rs7153397 with CRISPR interference (CRISPRi) and analysed *CCDC88C* expression in breast cancer cohorts.

### CCV selection and coordinate conversion

We selected 5,116 CCVs reported by BCAC ^1^, excluding 2,277 in strong LD with a copy number variant (index variant rs62070949) and one withdrawn from the hg38 version of the human reference genome (rs72127681).

We converted the genomic coordinates of CCVs from human genome build hg19 to hg38 using SNP rs IDs as input against dbSNP version 151 and dbSNP version 155 using BCFtools version 1.11 ‘view’ function, within R 3.6.0 environment. The hg38-equivalent IDs and coordinates for COSMIC variants and historically merged SNPs were inputted manually (rs1491361031, rs36094352, rs78105048, rs146915990, rs587714804, rs143207801, rs114312440, rs559449702, rs142901261, rs1353154669, rs533235186, rs111769614, rs528673091, rs8039, rs142118767, COSM3756504). Where a variant did not have an associated rs ID in the BCAC hg19 list, position was queried using liftOver version 20180306.

### Oligonucleotide design

We designed 20,878 oligonucleotides of 270 bp centred on reference (REF) or alternative (ALT) alleles in forward and reverse orientations, with 15 bp adaptors^3^. We extracted the up-and down-stream nucleotide sequences surrounding each CCV in the hg38 human reference genome, using a custom R script. For SNPs, 135 bp downstream and 134 bp upstream of the CCV was selected, and for insertion/deletion polymorphisms, flanking sequence length was adjusted depending on variant length, such that flanking sequence remained consistent for each pair of reference and alternative alleles. Adaptors were added in both orientations to create forward and reverse sequences for each CCV. The library included 36 positive controls near *GATA3* and *PGR* and 400 negative controls from scrambled sequences. Scrambled negative control sequences were generated by randomly sampling nucleotides to match the length of test sequences, excluding homopolymers and the SceI restriction enzyme recognition site. Each negative sequence was paired with a single-nucleotide variant at a fixed position to mimic REF and ALT alleles. Oligonucleotides were synthesised by Twist Bioscience.

### Library generation

The oligonucleotide pool was amplified via two-stage PCR. A 5-cycle PCR added minimal promoter and adaptor sequences downstream of the CRS, and the second 15-cycle PCR added a 15 bp random barcode sequence downstream of the first-round fragment. Amplified sequences were ligated into a pLS-SceI vector (Addgene #137725^3^) using NEBuilder HiFi DNA Assembly Master Mix (NEB) and electroporated into 10-beta electrocompetent cells (NEB, C3020) using a Eppendorf Eporator. Colonies were grown overnight on ampicillin plates, supplemented with 100 µl/plate of 100mg/ml carbenicillin (Teknova). The plasmid library was purified with QIAgen Plasmid Plus Midi Kit (Qiagen, 12943). 3.13 million colonies were harvested to ensure each oligonucleotide sequence was associated with around 150 barcodes. The plasmid library was sequenced to determine barcode sequences and their CRS association. CRS-barcode fragments were amplified to add P5 and P7 flowcell sequences upstream and downstream, respectively. Sequencing was carried out using NextSeq P1 with 300 cycles and custom primers ^3^.

### Cell line culture

T-47D cells (HTB-133, ATCC) were cultured in RPMI-1640 (Gibco) supplemented with 10% foetal bovine serum (FBS; Gibco) and 10 μg/ml human insulin (Sigma) at 37°C, 5% CO_2_. 293T cells (CRL-3216, ATCC) were cultured in DMEM (Gibco) supplemented with 10% FBS at 37°C, 5% CO_2_. Cells were routinely tested for Mycoplasma contamination.

### Lentivirus packaging and titration

Lentivirus packaging and titration was carried out as described previously^3^. 293T cells were seeded at 1.1×10^7^ cells per T225 flask and incubated for 48 hours, four flasks were used for the 3 million barcode library. Cells were transfected with 10 μg/flask plasmid library, 6.5 μg/flask psPAX2 (Addgene #12260) and 3.5 μg/flask pMD2.G (Addgene #12259) using EndoFectin (GeneCopoeia) and incubated for 8.5 hours. Cell media was refreshed with 5% FBS and 1x ViralBoost reagent (Alstem) and cultured for a further 40 hours. Supernatant was filtered to isolate lentivirus and then concentrated using Lenti-X concentrator reagent (Takara).

T-47D cells were seeded for lentivirus titration at 1.75×10^5^ cells per well of a 24-well plate and incubated for 24 hours. Lentivirus (0, 1, 2, 4, 8, 16, 32 or 64 μl) was added with Polybrene (Sigma-Aldrich) at a final concentration of 8 μg/ml and cells were incubated for 24 hours. The next day fresh media was added without Polybrene, and the cells were incubated for a further 2 days. Cells were washed three times with DPBS, and genomic DNA was extracted using Wizard SV genomic DNA purification kit (Promega). Multiplicity of infection (MOI) was determined as the ratio of viral DNA to genomic DNA, accounting for relative plasmid backbone DNA, which represents non-integrated virus. This was measured by quantitative PCR (qPCR) using SYBR Green Mastermix (Applied Biosystems).

### Lentiviral infection and DNA and RNA sequencing

Lentiviral infection and subsequent DNA and RNA sequencing was carried out as described previously^3^ in triplicate, to obtain three biological replicates. T-47D cells were seeded at 3.7×10^6^ cells per replicate, to allow for 90 integrations per barcode, in a 15 cm plate and incubated for 24 hours. Cells were infected with the lentiviral library, at an estimated MOI of 80, and Polybrene (Sigma-Aldrich) was added at 8 μg/ml final concentration. The next day fresh media was added without Polybrene, and cells were cultured for two more days. Cells were washed three times with DPBS, and genomic DNA and total RNA were extracted from cells using the AllPrep DNA/RNA Mini Kit (Qiagen) and RNase-Free DNase Set (Qiagen). RNA samples were DNase treated, using TURBO DNA-free Kit (Invitrogen) and reverse transcribed to add a 16 bp UMI and P7 flowcell sequence downstream of the barcode. We continue to refer to these samples as RNA in subsequent steps to distinguish them from genomic DNA samples. Library prep was carried out following the protocol for a high-complexity library^3^, with a 3-cycle first round PCR reaction to add the P5 flowcell sequence and sample index sequence upstream and 16 bp UMI and P7 flowcell sequence downstream of the barcode. The second round PCR reaction was carried out with 17 cycles for DNA samples and 15 cycles for RNA samples. Sequencing was completed on a NovaSeq 6000 using custom primers.

### LentiMPRA data analysis

Sequences from both the association and DNA/RNA sequencing steps were output in fastq format. From the association step there were 108 million reads, with a length of 146 bp (CRS upstream and CRS downstream) or 15 bp (Barcode). The DNA/RNA step produced between 56.9 and 88.7 million reads per replicate for DNA, and between 144.8 and 283.6 million reads per replicate for RNA, with a length of 15 bp (barcode forward and barcode reverse) or 16 bp (UMI).

Raw fastq sequences from the association step were input into a custom shell script to add 15 bp adaptors to each end of the sequences. The association.nf function of MPRAflow^3^ was ran in Nextflow version 20.10.0 with the cigar flag specified as 300M and mapq flag set to 30, producing a pickle file as output. This output file was used to obtain barcode counts with MPRAflow Pipeline Version 2.3.5; CRS were removed if they were associated with less than 20 unique barcodes.

Raw fastq sequences from the DNA/RNA count step were ran with the count.nf function of MPRAflow^3^ using Nextflow version 20.10.0 with the flags thresh 20, bc-length 15, umi-length 16 and merge-intersect TRUE. The run was repeated with the --mpranalyze flag to produce input for MPRAnlayze^6^. This input was formatted using a custom script within R (v4.4.1) to exclude CRS with no barcodes, specify positive and negative controls, remove CRS if either DNA or RNA data was missing, and remove variants that did not have data for all four CRS types (i.e. fwd-REF, fwd-ALT, rev-REF, rev-ALT). Barcodes were limited to a maximum of 200 per CRS, and the resulting matrices were used to create an MPRAObject^6^. MPRAnalyze version 1.9.1 was used for this and subsequent MPRAnalyze analysis.

The library size of the MPRAObject was normalised using the estimateDepthFactors depth estimator totsum. Subsequent quantification analysis was carried out without parallelisation using the analyzeQuantification, getAlpha and testEmprirical functions of MPRAnalyze. The analyzeComparative function of MPRAnalyze was ran to compare differential allelic activity^6^.

### H3K27ac CUT&Tag data generation

T-47D cells were prepared for CUT&Tag following the protocol from Active Motif (https://www.activemotif.com/catalog/1320/cut-tag-service). Briefly, adherent cells were washed in 1X PBS and collected in 1X Enzyme Free Cell Dissociation Solution Hank’s Based (Sigma) using a cell scraper at room temperature. Cells were counted using a Countess II Automated Cell Counter (Thermo Fisher), pelleted at 500 xg for 5 minutes at 4°C, and resuspended in ice-cold preservation solution (50% FBS/40% growth media/10% DMSO (Sigma)) at a concentration of 400 cells/µl. 500 µl (200,000 cells) was transferred to a 1.5 ml tube on ice and cells were gradually frozen to -70°C in a styrofoam container. Cells were shipped on dry ice to Active Motif for H3K27ac CUT&Tag data generation. Briefly, cells were incubated overnight with concanavalin A beads and 1 µl of the primary anti-H3K27ac antibody per reaction (#39135, Active Motif). After incubation with the secondary anti-rabbit antibody (1:100), cells were washed and tagmentation was performed at 37°C using protein-A-Tn5. Tagmentation was halted by the addition of EDTA, SDS and proteinase K at 55°C, after which DNA extraction and ethanol purification was performed, followed by PCR amplification and barcoding (see Active Motif CUT&Tag kit, #53160 for recommended conditions and indexes). Following SPRI bead cleanup (Beckman Coulter), the resulting DNA libraries were quantified and sequenced on an Illumina NextSeq 550 (8 million reads, 38 bp PE reads).

### H3K27ac CUT&Tag data analysis

Reads were aligned using the BWA algorithm (mem mode; default settings)^15^. Duplicate reads were removed, and only reads that mapped uniquely (mapping quality ≥ 1) and as matched pairs were used for further analysis. Peaks were identified using the MACS 3.0.0 algorithm at a cutoff FDR corrected *P*-value of 0.05, without control file, and with the –nomodel option. Peaks that were on the ENCODE blacklist of known false ChIP-seq peaks were removed. Signal maps and peak locations were used as input data to Active Motif’s proprietary analysis program, which creates output containing information on sample comparison, peak locations and gene annotations. Consensus peaks per cell type were defined as the maximum coordinates of peaks present by intersect in both replicates.

### Oligonucleotide library annotation

We annotated the oligonucleotide library with available data using custom scripts in R (v4.4.1). We used direct intersect with Bedtools (v2.29.2) to annotate each CCV against gene promoters, H3K27ac peaks, DHS and TF ChIP-seq peaks. Gene promoters were defined as 1 kb downstream and 500 bp upstream of a transcription start site (TSS) in the Gencode v46 basic gene annotation table (UCSC table wgEncodeGencodeBasicV46^16^). T-47D TF ChIP-seq and DHS peaks were downloaded from ENCODE 4 hg38 as narrow peaks without further processing: ENCFF574HSR (*GATA3*), ENCFF420MLJ (*FOXA1*), ENCFF433NIE (*ESR1* treated with DMSO) ENCFF597QIF (*ESR1* treated with estradiol), ENCFF387ZTS (*EP300*), ENCFF177MVH (*JUND*) and ENCFF592NQU (DHS). CCVs were assigned to their nearest Gencode v46 basic TSS using R package: genomation (v1.38.0). Previously defined candidate target genes were also annotated from Fachal, et al. ^1^ (level 1 predicted target genes from Supplementary Table 6a; predicted target genes with largest confidence level for each variant from Table 1).

### CRISPR-based perturbation (CRISPRi)

Single guide RNAs (sgRNAs) were designed against target sequences for repression, using CHOPCHOP (http://chopchop.cbu.uib.no). Target sequences were selected based on their proximity to variants or gene promoters of interest, and specificity scores. Non-targeting scrambled sgRNAs from Gasperini, et al. ^17^ were used as negative controls (**Supplementary Table 4**). Complementary single-stranded DNA oligonucleotide pairs were designed with 5’-CACC (top strand) and 5’-AAAC (bottom strand) overhangs to facilitate cloning. If absent, a G nucleotide was substituted at the first base of the target sequence to improve transcription from the U6 promoter. DNA oligonucleotides were synthesised by IDT and cloning was performed as previously described^18^. Briefly, top and bottom strand sgRNA pairs were phosphorylated and annealed in a 10 µl PCR reaction, then diluted to 7.1 fmol/μl in EB Buffer (Qiagen). The guide expression vector pKLV2-U6gRNA5(BbsI)-PGKpuro2AZsG-W (Addgene #67975^19^) was linearised in a 50 µl reaction using 2 µg with 30 U BbsI (NEB) and 1x NEBuffer r2.1 (NEB). Annealed oligonucleotides were ligated into gel purified linear vector in a 10 µl reaction using 2 µl oligonucleotides, 20 ng linear vector, 20 U T4 ligase (NEB) and 1x ligase buffer (NEB), which was incubated at 16°C for a minimum of 5 hours. The ligation mixture was transformed using 5 µl with 50 µl chemically competent bacterial cells, and successful cloning was confirmed by Sanger sequencing (GENEWIZ, Azenta Life Sciences).

Cloned guide expression vectors were packaged into lentivirus in 293T cells. For each vector, 2×10^6^ 293T cells were seeded in a T25 flask and incubated overnight. Cells were transfected with 500 ng/flask plasmid, 500 ng/flask psPAX2 (Addgene #12260) and 150 ng/flask pMD2.G (Addgene #12259) using 4 µl Lipofectamine 3000 (ThermoFisher Scientific), 2.5 µl P3000 reagent (ThermoFisher Scientific) and 125 µl OptiMEM (ThermoFisher Scientific) and cultured for 24 hours. Cell media was refreshed and cells cultured for a further 48 hours. Culture media containing lentivirus particles was collected and stored at 4°C. Fresh media was added to the flasks and cells were cultured for a further 24 hours. A second aliquot of media containing lentivirus particles was collected and added to the first. This was centrifuged at 1600 xg for 10 minutes at 4°C to pellet cellular debris. Supernatant was filtered through Millex-HV 0.45 μM PVDF filters (Millipore), concentrated with Lenti-X concentrator reagent (Takara) for 30 minutes at 4°C and centrifuged at 1500 xg for 45 minutes at 4°C. The pellet was resuspended in 1/10 of the original volume using DPBS and stored at 4°C before use.

T-47D cells stabling expressing dCas9-KRAB, for CRISPR interference (CRISPRi), had previously been derived from parental cell lines in the lab, by transduction with lentivirus. Lentivirus was packaged as described above, using Lenti-dCas9-KRAB-blast (Addgene #89567^20^) rather than sgRNA containing vectors. Parental cell lines were seeded in a T75 flask, cultured to 50% confluency and 500 μl lentiviral supernatant was added to cells for 24 hours. Media was refreshed without lentivirus and cells cultured for a further 24 hours before 10 μg/ml blasticidin (InvivoGen) was added to select successfully transduced cells. Expression of dCas9-KRAB was confirmed by Western blot with antibody to Cas9 (**Supplementary Fig.5**).

T-47D-dCas9-KRAB cells were seeded into 12-well plates at a density of 100,000 cells per well and incubated for 24 hours. sgRNA lentivirus at 100 μl/well was added and cells cultured for 72 hours. Media was refreshed with 1 μg/ml puromycin (InvivoGen) to select successfully transduced cells expressing Zsgreen (visualised by fluorescent microscope). Cells were collected and RNA extracted using the RNeasy Kit (Qiagen). RNA was converted to cDNA using 2 μg with 1 µl 500 µg/ml oligo (dT)_15_ primer (Promega), 1 µl 10 mM dNTP mix (Invitrogen), 4 µl 5X First Strand Buffer (Invitrogen), 1 µl 0.1 M DTT (Invitrogen), 1 µl RNase-OUT (40 U/µl) (Invitrogen) and 1 µl 200 U/µl SuperScript III Reverse Transcriptase (ThermoFisher Scientific), according to manufacturer’s protocol.

### qPCR

Gene expression in CRISPRi cells transduced with sgRNA lentiviral particles was measured using TaqMan assays specific to the gene of interest. A 5 μl reaction was prepared with 1:10 dilution of cDNA, 2.5 μl TaqMan Universal Master Mix II (no UNG, Applied Biosystems) and 0.25 μl 20X gene-specific TaqMan Assay (*CCDC88C* (Hs00380245_m1), *C14orf159*Hs01061067_m1), *PPP4R3A* (Hs00215697_m1), *GAPDH* (Hs00266705_g1), all Applied Biosystems). Amplification was measured on QuantStudio v. 6 Flex Real-Time PCR System. Relative gene expression, compared to empty vector alone and normalised to *GAPDH*, was calculated using the ΔΔCT method.

### Western blotting

Cells were washed in PBS and whole cell extracts were prepared by lysing cells with lysis buffer (150mM NaCl, 50mM Tris pH8.0, 1% Triton X-100, PhosStop and Complete Mini EDTA free protease inhibitors (Roche)) on ice for 5 minutes. Lysed cell suspension was collected using a cell scraper and agitated for 30 minutes at 4°C. Following centrifugation at 13,000 rpm for 25 minutes at 4°C, the whole cell lysate supernatant was aliquoted and stored at -70°C. Protein yield was quantified by Bio-Rad Protein Assay Dye Reagent Concentrate (Bio-Rad) with BSA (NEB) as standard. Whole cell lysates were subjected to SDS-PAGE on a 4-12% Bis Tris gel with MOPS buffer (Invitrogen) and transferred to a nitrocellulose membrane (0.45 µm pore size, Invitrogen). The membrane was blocked with 5% non-fat milk in TBST (1X TBS, 0.05% Tween 20) at room temperature overnight with shaking. The membrane was probed with an antibody against dCas9 ((7A9-3A3) N-Terminus-BSA Free, NBP2-36440, Novus Biologicals) diluted 1:1,000; or beta-actin (as loading control; AC-15, monoclonal, Sigma A5441) diluted 1:40,000; in 5% non-fat milk in TBST for 2 hours at room temperature. After washing 3 times with TBST, the blots were incubated with anti-mouse IgG secondary antibody (A9044, Sigma) or anti-mouse IgG (H+L) (115-035-003, Jackson Immuno Research) conjugated to horseradish peroxidase diluted 1:10,000 in TBST for 1 hour at room temperature. The blot was then washed four times with TBST and developed with ECL Detection Reagent (Bio-Rad) (**Supplementary Fig.5**).

### Data analysis

The Cancer Genome Atlas (TCGA) data were downloaded from the GDAC portal at: http://gdac.broadinstitute.org (release 28-January-2016) and SCAN-B from Gene Expression Omnibus (GEO), with accession ID GSE96058. The datasets were pre-processed as described previously^21,22^. Differential expression comparison of *CCDC88C* between breast cancer subtypes ER+ and ER-, and normal breast tissue for TCGA and SCAN-B, was performed using the Welch’s t-test in R (v4.4.2). Survival analysis was performed using the Cox proportional hazards model in R (v4.4.2) with *P*-values estimated using the Wald-test, R package: survival (v3.8-3). Locus plots were created in R (v4.4.1) using Gviz (v1.50.0) with custom scripts. Graphs were produced in Prism (v10.4.1). The position weighted matrix for the *FOXA1* binding site was downloaded from Rauluseviciute, et al. ^23^. Correlation figures of normalized log_2_ DNA counts and RNA counts were replicated from MPRAflow^3^. Summary figure generation and custom post-hoc editing were performed in Adobe Illustrator v29.6.1. T-47D DNase-seq profiles were plotted from bigwig output downloaded from ENCODE 4 hg38 ENCFF085GPE. QQ plots were generated in R (v4.2.1) using the inbuilt qqnorm and qqplot R functions.

### Statistical analysis

#### LentiMPRA

We obtained MPRAnalyze output in the format of alpha with p-values for MAD-score (a median-based variant of z-score), allele log_2_FC with FDR-corrected *P*-values and direction log_2_FC with FDR-corrected *P*-values. We used these as estimates of enhancer activity, functionality of variant and evidence of directionality of enhancer, respectively. For all measures we used an FDR corrected *P*-value with a threshold of 0.1. When FDR corrected *P*-values were not produced by MPRAnalyze, these were calculated in R (v4.4.1). For the stratified analysis to estimate allelic log_2_FC on forward and reverse strands separately, we plotted the quantiles of log_2_FC on the forward strand against the quantiles of log_2_FC on the reverse strand.

#### Enrichment analysis

We stratified CRS on regulatory activity (significant = alpha *P* < 0.1; non-significant = alpha *P* > 0.1) and categorised them based on absolute allele fold change. We carried out a Pearson’s chi-squared test on these categories in R (v4.4.1). We carried out a two-sided Fisher’s exact test in R (v4.4.1) to determine annotations with significant enrichment in differentially expressed variants, using a Bonferroni corrected *P*-value of 0.006 (8 tests).

#### qPCR

For the negative control sgRNAs (Scrambled_1 and Scrambled_2), we used t-tests to test H0: the relative gene expression does not differ from 1.0. To maximise the power of subsequent analyses, we then combined the negative control data and for each of the other sgRNAs we tested H0: relative gene expression does not differ from the combined negative control relative gene expression. To account for multiple testing, we used a Bonferroni corrected p value of 0.006 (8 tests per gene).

## SUPPLEMENTARY FIGURES

**Supplementary Figure 1.**
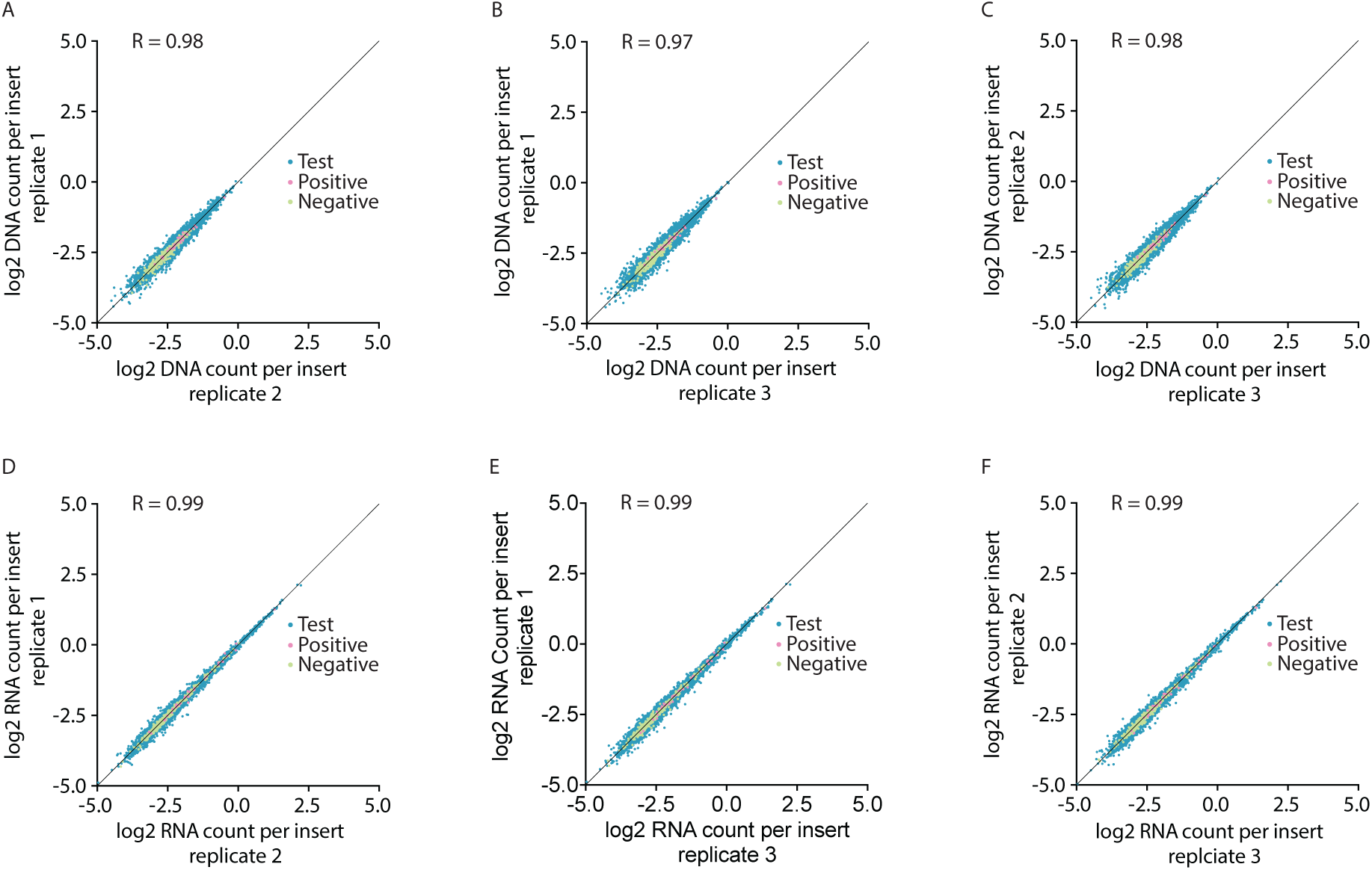
Replicate correlations. **A-C,** Correlation of log_2_ normalized DNA count per insert of **(A)** replicate 1 against replicate 2, **(B)** replicate 1 against replicate 3, **(C)** replicate 2 against replicate 3. **D-F,** Correlation of log_2_ normalized RNA count per insert of **(D)** replicate 1 against replicate 2, **(E)** replicate 1 against replicate 3, **(F)** replicate 2 against replicate 3.

**Supplementary Figure 2.**
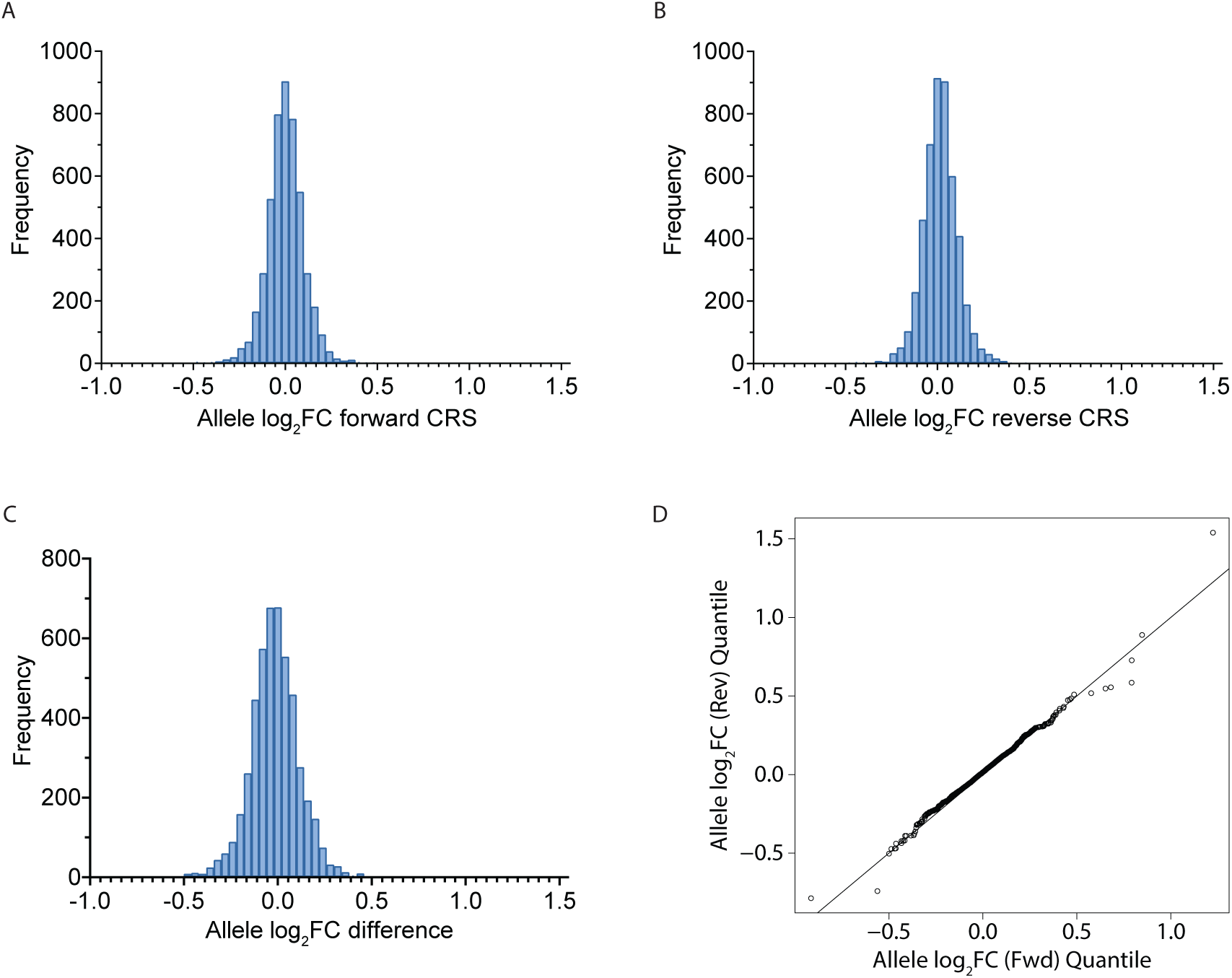
Allelic log_2_FC analysis. **A,B,** Frequency of allele log_2_FC from stratified analysis testing **(A)** forward CRS and **(B)** reverse CRS independently. **C,** Frequency of the diEerence between output from stratified analysis of allele log_2_FC on the forward strand and allele log_2_FC on the reverse strand. **D,** QQ plot of the quantiles of allele log_2_FC on the forward strand against quantiles of allele log_2_FC on the reverse strand.

**Supplementary Figure 3.**
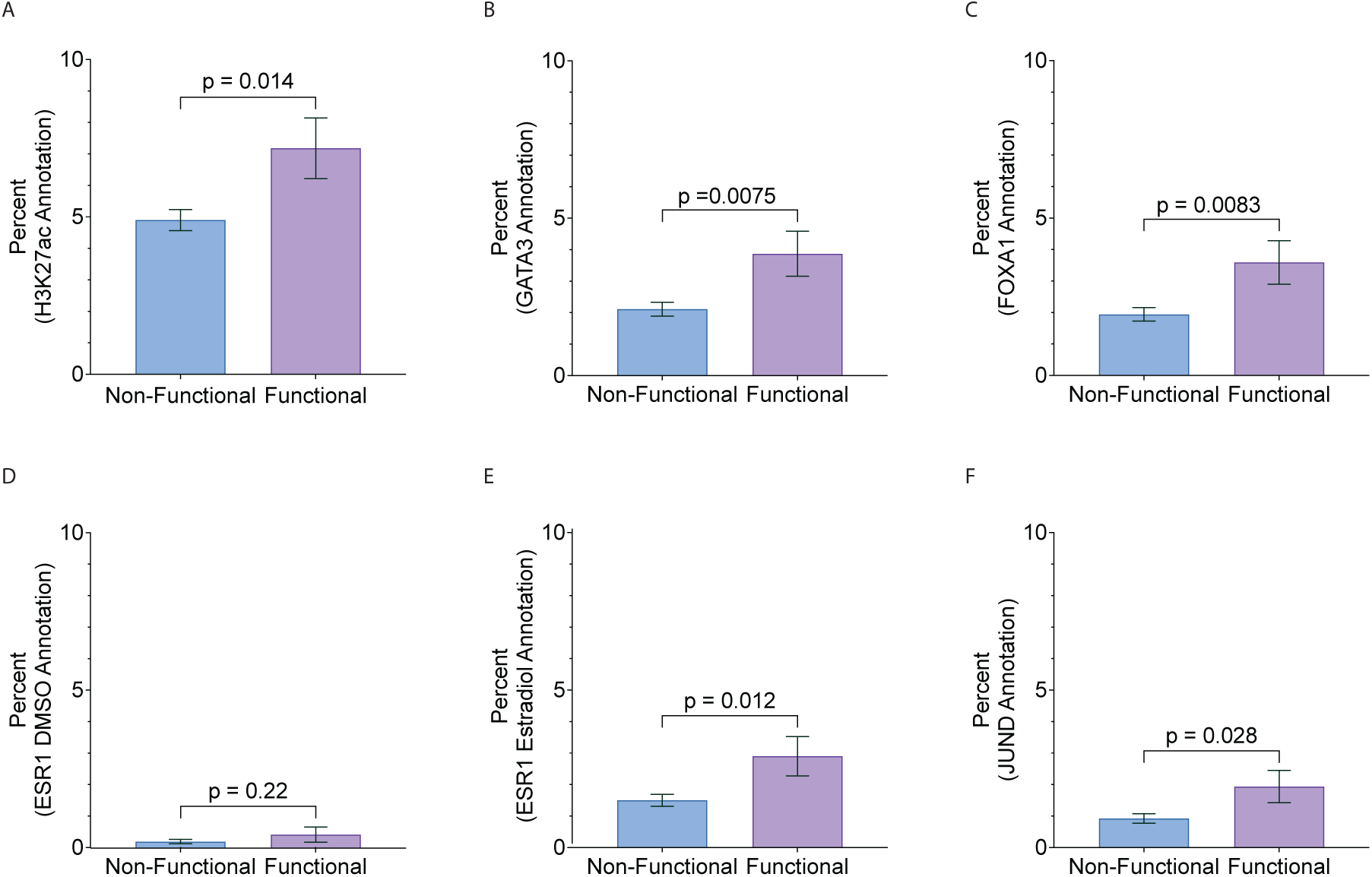
Additional enrichment. **A-F,** Percentage of sequences with H3K27ac **(A),** *GATA3* **(B)** *FOXA1* **(C),** *ESR1* (DMSO) **(D)** *ESR1* (estradiol) **(E)** and *JUND* **(F)** annotations comparing non-functional (allele log_2_FC *P* > 0.1) and functional variants (allele log_2_FC *P* < 0.1). Error bars denote standard error of the mean; *P* values from two-sided Fisher’s exact test, and a Bonferroni correction was applied to account for multiple testing with a *P* value of 0.006 considered as significant. All non-significant.

**Supplementary Figure 4.**
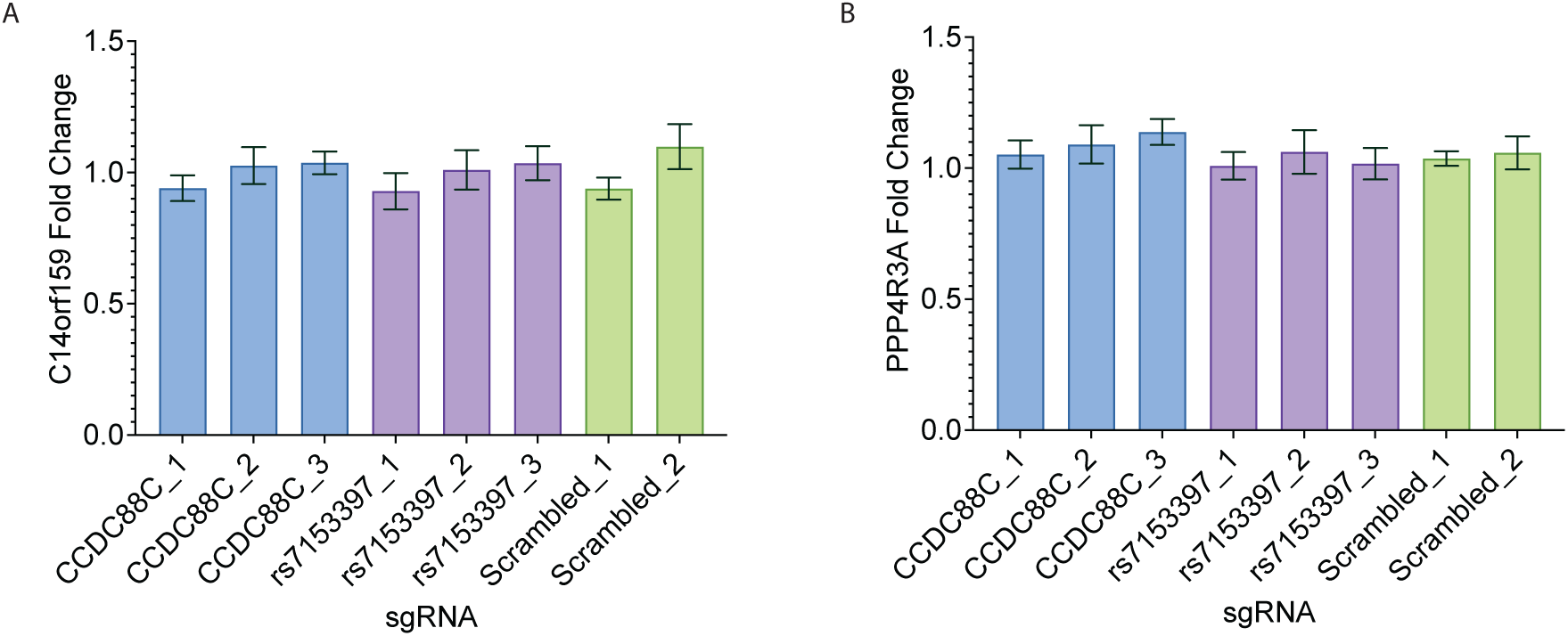
CRISPRi gene expression. **A,B,** Expression levels of *C14orf159* **(A)** and *PPP4R3A* **(B)** in T-47D cells following CRISPRi with sgRNAs targeting dCas9-KRAB to the *CCDC88C* promoter (CCDC88C_1, CCDC88C_2, CCDC88C_3), rs7153397 variant (rs7153397_1, rs7153397_2, rs7153397_3) or non-targeting negative controls (Scrambled_1, Scrambled_2). Fold change was corrected for *GAPDH* and relative expression (compared to empty vector alone) was calculated for each sgRNA using the ΔΔCT method. Error bars denote standard error of the mean; *P* values from *t*-tests and a Bonferroni correction was applied to account for multiple testing with a *P* value of 0.006 considered as significant. All non-significant.

**Supplementary Figure 5.**
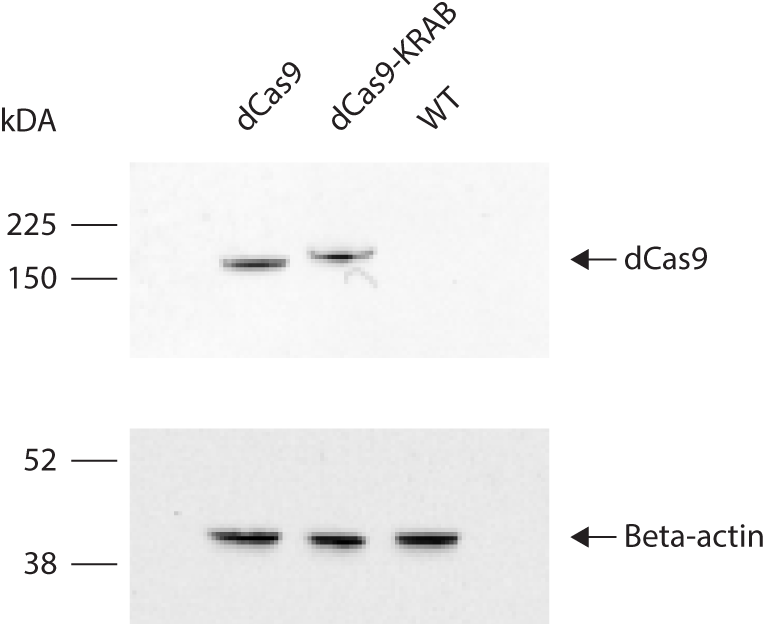
dCas9 western blot. Western blot of whole cell lysates from T-47D cells that have been modified to express dCas9 (lane 1, positive control), dCas9-KRAB (lane 2) and T-47D wildtype cells (lane 3, negative control). Blots were probed with an anti-dCas9 antibody. As a loading control, blots were probed with an antibody against beta-actin.

## Data Availability

All data produced in the present work are contained in the manuscript

## ACKNOWLEDGEMENTS

We thank the breast cancer patients who contributed to TCGA and SCAN-B by generously providing breast tissue samples and data. We thank the Breast Cancer Now Toby Robins Research Centre Data Science team for bioinformatics support and thank Breast Cancer Now for supporting the work of this team. The results shown here are in whole or part based on data generated by the TCGA research network: https://www.cancer.gov/tcga.

pLS-SceI was a gift from Nadav Ahituv (Addgene plasmid # 137725; http://n2t.net/addgene:137725; RRID:Addgene_137725) psPAX2 was a gift from Didier Trono (Addgene plasmid # 12260; http://n2t.net/addgene:12260; RRID:Addgene_12260)

pMD2.G was a gift from Didier Trono (Addgene plasmid # 12259;http://n2t.net/addgene:12259; RRID:Addgene_12259)

pKLV2-U6gRNA5(BbsI)-PGKpuro2AZsG-W was a gift from Kosuke Yusa (Addgene plasmid # 67975; http://n2t.net/addgene:67975; RRID:Addgene_67975)

Lenti-dCas9-KRAB-blast was a gift from Gary Hon (Addgene plasmid # 89567; http://n2t.net/addgene:89567; RRID:Addgene_89567)

## Author contributions

Conceptualization: OF, SH, NJ

Methodology: KM, OF, SH, NJ

Investigation: KM, HK, AG, KT, SS

Formal analysis: KM, HK, OF, SH, YL, JBS, MW, PL, RSH, NO

Supervision: OF, SH, NJ

Writing-original draft: KM, OF, RSH, HK, SH, NJ

Writing-review and editing: all authors

**Authors declare that they have no competing interests.**

